# A comparative study of real-time RT-PCR based SARS-CoV-2 detection methods and its application to human derived and surface swabbed material

**DOI:** 10.1101/2020.11.23.20236257

**Authors:** Aizhan Tastanova, Corinne Isabelle Stoffel, Andreas Dzung, Phil Fang Cheng, Elisa Bellini, Pål Johansen, Agathe Duda, Stephan Nobbe, Reto Lienhard, Philipp Peter Bosshard, Mitchell Paul Levesque

**Author notes:** Correspondence to: Mitchell Paul Levesque, University of Zurich, University Hospital Zurich, Department of Dermatology, Wagistrasse 14, CH- 8952, Schlieren, Zürich, Switzerland. These authors contributed equally (Aizhan Tastanova, Corinne Isabelle Stoffel, Andreas Dzung, Phil Fang Cheng, Philipp Peter Bosshard). – Lead author (Mitchell Paul Levesque).

## Abstract

Real-time reverse transcription polymerase chain reaction (RT-PCR) remains a gold standard in detection of various viral diseases. In the COVID-19 pandemic, multiple RT-PCR based tests were developed to screen for viral infection. As an emergency response to growing testing demand, we established a SARS-CoV-2 PCR diagnostics platform for which we compared different commercial and in-house RT-PCR protocols. We evaluated four commercial (CDC 2019-nCoV, Applied Biosystems™ 2019-nCoV Assay Kit v1 TF-SinglePlex, 2019-nCoV Assay Kit v2 TF-MultiPlex, and EURORealTime SARS-CoV-2), one customized (Institute Pasteur), and one in-house RT-PCR protocols with 92 SARS-CoV-2 positive and 92 SARS-CoV-2 negative samples. Furthermore, we compared economical and practical characteristics of these protocols. We also developed a highly sensitive digital droplet PCR (ddPCR) method. Finally, we conducted a local environmental study for the presence and infectivity of SARS-CoV-2 on different surfaces in a quarantined household using RT- and ddPCR methods. We found very low limits of detection (1 or 2 viral copies/μL), high sensitivities (93.6-97.8%) and specificities (98.7-100%) for the tested RT-PCR protocols. We further demonstrated the feasibility of downscaling two of the commercial protocols, which could optimize testing capacity. In the local environmental study, only one surface sample tested positive for viral RNA, but without detectable infectivity in vitro. Tested commercial and customized RT-PCR detection kits show very good and comparable sensitivity, and specificity, and the kits could be further optimized for use on SARS-CoV-2 viral samples derived from human and surface swabbed samples.

## Introduction

On March 11, 2020, the World Health Organization (WHO) declared a pandemic due to the quick spread of a respiratory disease caused by the novel Severe Acute Respiratory Syndrome Coronavirus 2 (SARS-CoV-2). With positive cases growing in multiple countries and due to its high transmissibility, eradicating SARS-CoV-2 is rather unrealistic in the short term (1). In Switzerland, the second wave of the SARS-CoV-2 is predicted to be slower than the first one, but with a higher case fatality rate (2). The same situation was reported for the Spanish Flu by the WHO, where the 2nd and 3rd waves of the infection claimed more lives and the pandemic lasted for almost 2 years and resulted in at least 50 million deaths Worldwide (3). Another important factor contributing to the rapid spread of COVID-19 pandemic is an unusually high number of asymptomatic spreaders (4, 5). Therefore, continuous testing and reliable detection of the virus is an essential part of controlling the spread of SARS-CoV-2 (6).

In March 2020, we initiated an in-house platform for SARS-CoV-2 diagnostics in a routine microbiology lab as part of an emergency response to a growing demand for test capacity. Currently, the gold standard for the detection and diagnosis of SARS-CoV-2 infection is based on the real-time reverse transcription PCR (RT-PCR). Our overall goal was to provide in-house SARS-CoV-2 diagnosis to all patients and personnel, to ensure the safe and efficient continuation of the health care work within the hospital and the protection of high-risk patients. The aims of this study were: *i)* to evaluate four commercially available, one customized, and one in-house RT-PCR test by comparing the limit of detection (LoD), sensitivity using a panel of SARS-CoV-2 confirmed cases, and specificity using a group of non-COVID respiratory samples; *ii)* to examine the feasibility of down-scaling two commercial protocols in order to optimize the testing capacity; *iii)* to develop a droplet digital PCR (ddPCR) assay to increase test sensitivity and provide more accurate quantitation of viral RNA; and *iv)* to conduct a local environmental study in a quarantined household by using two validated RT-PCR and ddPCR protocols.

## Methods

### Clinical samples

Patient samples were collected by nasopharyngeal and/or oropharyngeal swabs (iClean, CM-FS913, San Ramon, USA) at the University Hospital Zurich and at ADMed Laboratory in La Chaux-de-Fonds (COPAN, Brescia, Italy). The non-COVID-19 samples (other respiratory disease samples) were kindly provided by ADMed Laboratory and were selected after having been tested on the Respiratory Panel FilmArray on Biofire (bioMérieux). Household samples were collected by swabbing of the different surfaces in a quarantined household of a SARS-CoV-2 positive patient. All swabs were stored in a viral transport medium (CDC formulation (7) or Eswab (AMIES, COPAN, Murrieta, United States)) at 4 °C for max. 48 hours or stored at -80 °C until further analyses. All household swabbing participants provided informed consent for the study and both the assay establishment and household studies were approved by the Cantonal Ethics Committee: BASEC-Nr-2020-00660 and BASEC-Nr-2020-00659, respectively.

### RNA extraction

Viral RNA was extracted as previously described using magnetic bead-based (SpeedBeads, GE Healthcare, Darmstadt, Germany) extraction kit for KingFisher instrument (MagMax, Thermo Fisher, Waltham, Massachusetts, U.S) (8).

### Detection of SARS-CoV-2 by Reverse-Transcriptase PCR (RT-PCR) protocols

Four commercially available, one customized from Pasteur Institute and in-house optimized RT-PCR protocols (description available in S1 Table) were compared. Primer probes design, reaction mix and thermal cycling conditions are shown in S2, S3 and S4 Tables, respectively. All RT-PCR protocols were run according to manufacturer instructions on a QuantStudio 5 real-time PCR-System (Applied Biosystems, Switzerland) and data were analyzed with the Design and Analysis Software DA 2.4 (Applied Biosystems) except for the Euroimmun protocol, which was run on LightCycler® 480 II (RocheDiagnostics, Switzerland). Fast cycling mode was used and a comparative Ct analysis method was performed.

For the CDC protocol, a RT-PCR result was defined as *inconclusive* if only the N1 gene (plus/minus N3 gene) was positive, or if only the N2 gene (plus/minus N3 gene) was positive. For the TF-MultiPlex, TF-SinglePlex, and Oncobit protocols, a RT-PCR result was considered as *inconclusive* if only one out of 2 or 3 of the viral genes was positive. Inconclusive results were not repeated. The Euroimmun protocol does not have the *inconclusive* category.

### Detection of SARS-CoV-2 by Droplet Digital PCR (ddPCR)

The droplet digital PCR (ddPCR) protocol for SARS-CoV-2 detection targets two viral genomic regions of the SARS-CoV-2 gene (ORF1ab and N2) and uses the human RNase P gene as an in-process control. We used the following probes for the three genes: ORF1ab (FAM and HEX), N2 (FAM) and RNase P (HEX). Briefly, 20 µL of reaction mix (containing 1-Step RT-ddPCR Advanced Kit for Probes Mastermix (1864022, Bio-Rad)) was combined with 10µL of RNA sample for a final reaction volume of 30 µL. Final concentration of primers (ORF1ab, N2, RNaseP) was 90 nM, RdRP probes was 19.5 nM, N2 probe was 30 nM and RNase P probe was 40nM. The SARS-CoV-2 Positive Run Control (#COV019CE, Bio-Rad) was used as positive control. DdPCR was run according to the program shown in S5 Table using QX200 Droplet Digital PCR System (Bio-Rad). The swabbing household samples from laptop, newspaper, door handle, as well as the non-template control (NTC) were tested in two independent runs.

### Limit of detection, sensitivity and specificity calculation

Limit of detection (LoD) of four published SARS-CoV-2 detection protocols (CDC, TF-MultiPlex, TF-SinglePlex, and Euroimmun) was determined by using a dilution of an external quality assessment (EQA) quantitative test sample (9) Linear regression was used to determine the line of best fit for relationship between Ct and viral copies. A Ct value of 40 was set to be the minimum amount of viral copies detected by RT-PCR. LoD for Oncobit ddPCR protocol was determined by using a dilution of the SARS-CoV-2 Positive Run Control (#COV019CE, Bio-Rad).

For sensitivity and specificity value calculations of each assay, the results of RT-PCR obtained from the ADMed Laboratory (La Chaux-de-Fonds) were used as gold standard reference. The sensitivity was defined with the following formula “TP/(TP+FN)”, while specificity was defined as “TN/(TP+FP)”where: TP is true positive, FP is false positive, TN is true negative, and FN is false negative. If the result of tested assays matched the reference, it was labeled concordant. If the result from the tested assays did not match the gold reference, it was labeled discordant. Inconclusive results were excluded from sensitivity and specificity calculations.

### SARS-CoV-2 infectivity assay

The viral infectivity assay was performed as previously described (10-12) with slight modifications. Briefly, 5×10^4^ Vero E6 cells (ATCC #CCL-81) were seeded on 96-well flat bottom cell culture plates in 200 uL of high glucose DMEM medium supplemented with L-Glutamate, sodium pyruvate, non-essential amino acids, HEPES, 5% fetal cow serum (FCS), and Normocin^™^ (Cat. No. ant-nr-1, InvivoGen, Toulouse France). After 24 hours incubation (37 °C, 5% CO_2_), the medium was removed and 100 µL virus test solution or the positive SARS-CoV-2 control (kindly provided by Prof. Volker Thiel, Inst. Virology & Immunology, University of Berne) was added in 2-fold serial dilutions to the cells. The plates were incubated for 48 hours at 37 °C. The cells were then fixed with 10% formaldehyde solution for 15 minutes at room temperature, rinsed with phosphate buffered saline (PBS), and stained with 1% crystal violet stain solution (Pan Reac AppliChem Cat. No. 252532.1211, Darmstadt, Germany) for 15 minutes at room temperature. The staining solution was removed, the cells were rinsed twice with PBS, and the plates dried at room temperature before assessment for viral plaques.

## Results

### Description and comparison of SARS-CoV-2 RT-PCR detection protocols

The six RT-PCR protocols compared in this study utilize the same principle of isolating viral RNA from the nasopharyngeal and/or oropharyngeal swabs or bronchial fluid and running a 1-step reverse transcription reaction followed by real time amplification of two or three SARS-CoV-2 target genes (Figure 1A). We summarized and compared all tested RT-PCR protocols in Figure 1B. All protocols have internal (IC), non-template (NTC) and positive controls (PC). In TF-MultiPlex, the phage MS2 is added as IC that serves as both RNA isolation and reaction control. All other protocols except for Euroimmun (where type of IC is not indicated) use a widely accepted reaction control RNAseP to ensure that RNA isolation worked and RT-PCR reaction was not inhibited. Protocol’s design are either singleplex, doubleplex or multiplex. Euroimmun protocol stands out with its design, with two target probes coupled to the same reporter color FAM. The viral RNA input is 5 μL to 10μL.

**Figure 1:**
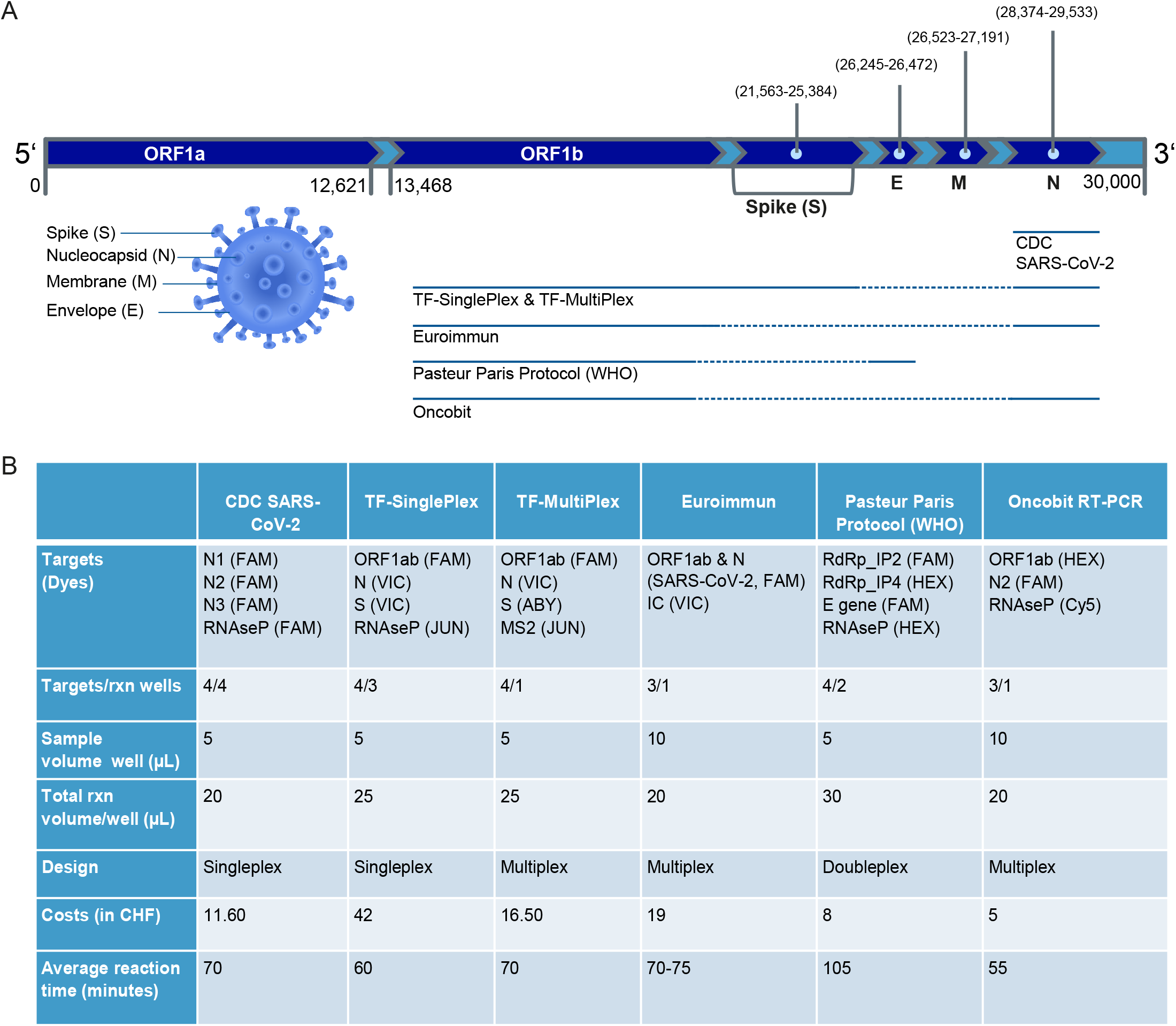
Summary of different SARS-CoV-2 real-time RT-PCR detection protocols. **A**. SARS-CoV-2 genome structure and coverage by different protocols, continuous line indicate relative gene coverage by the detection protocol. **B**. Comparative overview of six different RT-PCR protocols. *Abbreviations:* ***CDC SARS-CoV-2*** *(Centers for disease control and prevention SARS-CoV-2);* ***TF-SinglePlex*** *(Applied Biosystems™ TaqMan™ 2019-nCoV Assay Kit v1)*, ***TF-MultiPlex*** (Applied Biosystems™ Multiplex TaqMan 2019-nCoV Assay Kit v2 RESEARCH USE ONLY (R.U.O.) kit); ***Euroimmun*** (EURORealTime SARS-CoV-2 (For research use only (RUO)) *(Cat. No. MP 2606-0425));* ***Pasteur Paris Protocol (WHO)*** (Real-time RT-PCR assays for the detection of SARS-CoV-2, Institute Pasteur (Paris)); ***Oncobit real-time RT-PCR*** *(in-house customized RT-PCR protocol)*.

Due to unspecific E-gene amplification (S6 Table), the protocol developed by Pasteur institute in Paris was not used further in this comparative study.

### Limit of detection of real-time RT-PCR and ddPCR SARS-CoV-2 detection protocols

With a Ct-value cut-off of 40, the five RT-PCR SARS-CoV-2 detection protocols (CDC, TF-MultiPlex, TF-SinglePlex, Euroimmun, Oncobit) as well as Oncobit ddPCR protocol showed limit of detection (LoD) between 1 and 2 viral copies/μL (Figure 2A, B). Values below 1 copy/μL indicate high sensitivity of the tested protocol (Figure 2A, B).

**Figure 2:**
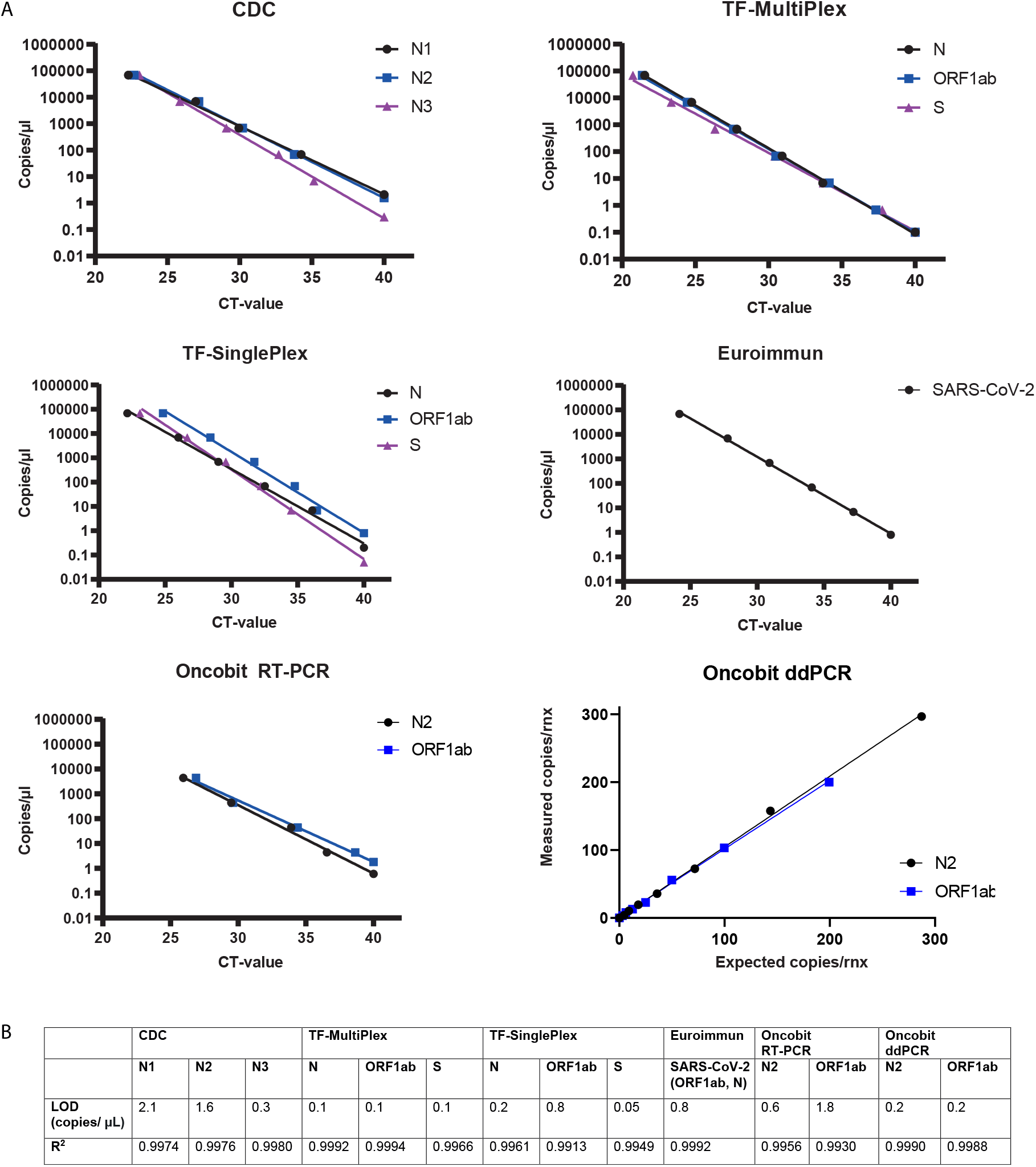
Limit of detection (LoD) of real-time RT-PCR and ddPCR SARS-CoV-2 detection protocols. **A**. Limit of detection (viral copies/µL) of different target genes of CDC, TF-SinglePlex, TF-MultiPlex, Euroimmun, Oncobit RT-PCR and Oncobit ddPCR SARS-CoV-2 detection protocols. **B**. Calculated R^2^ values of SARS-CoV-2 detection protocols.

### Specificity & Sensitivity of real-time RT-PCR SARS-CoV-2 detection protocols

For the sensitivity and specificity of the SARS-CoV-2 detection protocols (CDC, TF-SinglePlex, TF-MultiPlex, Euroimmun, Oncobit) we used a cohort of 92 SARS-CoV-2 positive samples and 92 SARS-CoV-2 negative samples that were kindly provided by ADMed Laboratory. A comparison to SARS-CoV-2 positive results showed similar sensitivity of all tested protocols with 93.6% for TF-SinglePlex and 96.7-97.8% for the other protocols (Figure 3A). In the specificity cohort, 22 samples had confirmed diagnosis of other respiratory diseases (S7 Table) and 70 samples tested negative for all listed respiratory diseases including SARS-CoV-2. All protocols, except TF-SinglePlex, showed no cross reactivity (Figure 3A) including samples that were positive for four other types of corona viruses (S7 Table). The specificity was thus 100% for all protocols except for TF-SinglePlex, which had a specificity of 98.7% (Figure 3A).

**Figure 3:**
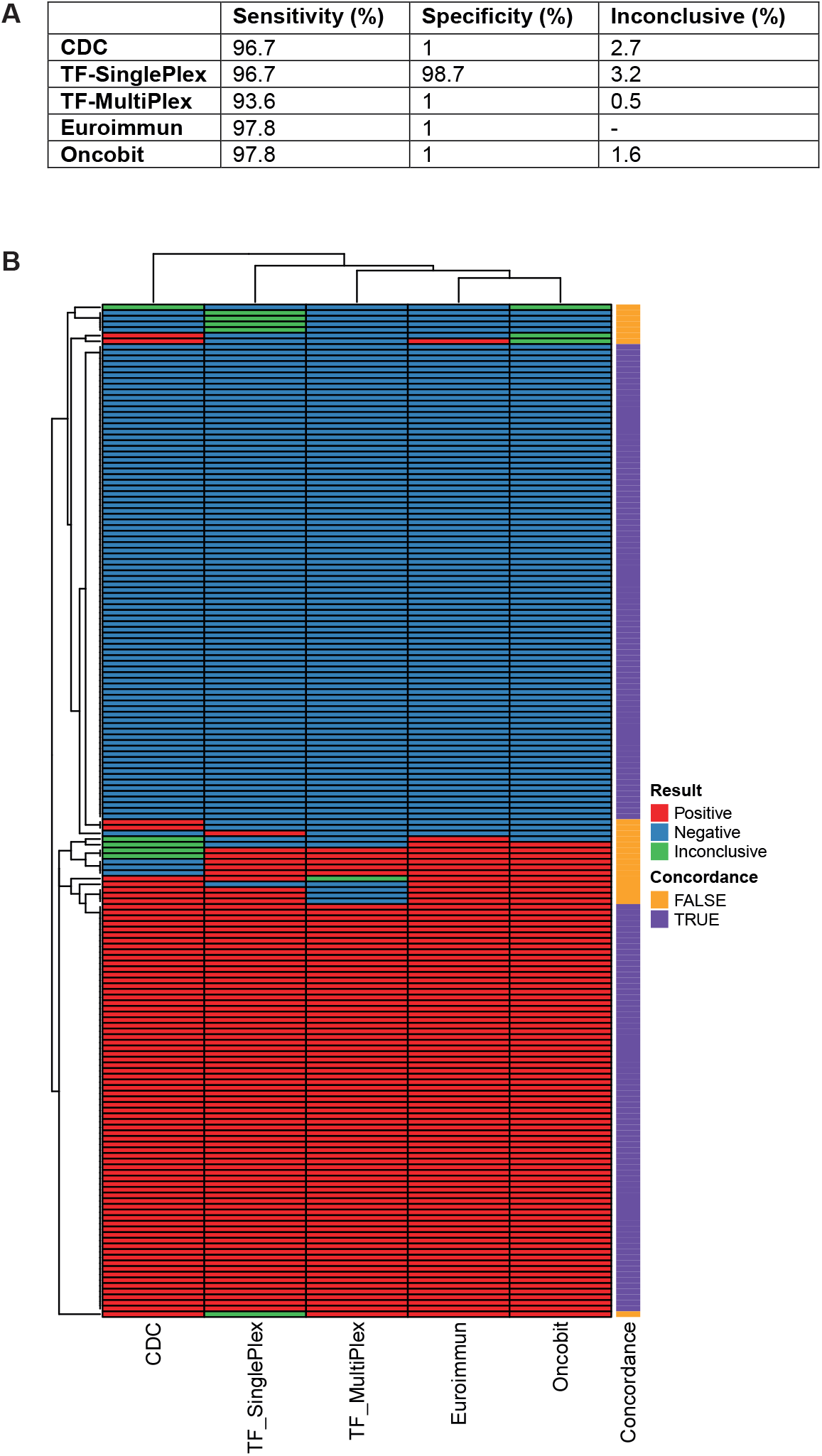
Specificity & Sensitivity of real-time RT-PCR SARS-CoV-2 detection protocols. **A**. Performance calculation (Sensitivity/Specificity) as well as calculation of percentage of inconclusive results of five real-time RT-PCR detection protocols (CDC, TF-SinglePlex, TF-MultiPlex, Euroimmun and Oncobit). Euroimmun RT-PCR detection protocol does not have the *inconclusive* category, *inconclusive* for Euroimmun equals *invalid* result. **B**. Heat-Map summarizing concordance of five real-time RT-PCR detection protocols (CDC, TF-SinglePlex, TF-MultiPlex, Euroimmun and Oncobit) for both sensitivity (bottom) and specificity (top) sample cohorts.

Inconclusive results were found in 0.5-3.2% of these 184 samples, with TF-MultiPlex and Oncobit providing the most accuracy (Figure 3A). Comparing RT-PCR results (positive, negative, inconclusive) of all 184 samples, we determined the overall non-concordance between all the protocols to be 14.7% (Figure 3B).

### Optimization of testing capacity

To optimize testing capacity, (i) we downscaled recommended reaction volumes in commercial protocols and (ii) we customized published primer/probe sequences to have an in house developed protocol (Oncobit). Using a previously confirmed SARS-CoV-2 positive cohort of 14 samples, we compared CDC and TF-MultiPlex protocols with recommended reaction volume and with reaction volumes reduced by 50%. RNA sample input was always the same. The Oncobit protocol was the cheapest (Figure 1B), had the shortest RT-PCR reaction time requirement (Figure 1B) and had most reliable access to consumables (Microsynth). The specificity and sensitivity of the Oncobit protocol was comparable to other commercial SARS-CoV-2 RT-PCR detection kits (Figure 3A,B).

A downscaled CDC protocol showed two (14.3%) *inconclusive* results, a standard TF-MultiPlex protocol showed one (7.1%) false negative result, and a downscaled TF-MultiPlex protocol revealed one (7.1%) false negative as well as one (7.1%) *inconclusive* result (Figure 4A). Furthermore, LoD of downscaled CDC and TF-MultiPlex protocols showed a sensitivity of 1 copy/μL with a Ct-value cut-off of 40 (Figure 4B).

**Figure 4:**
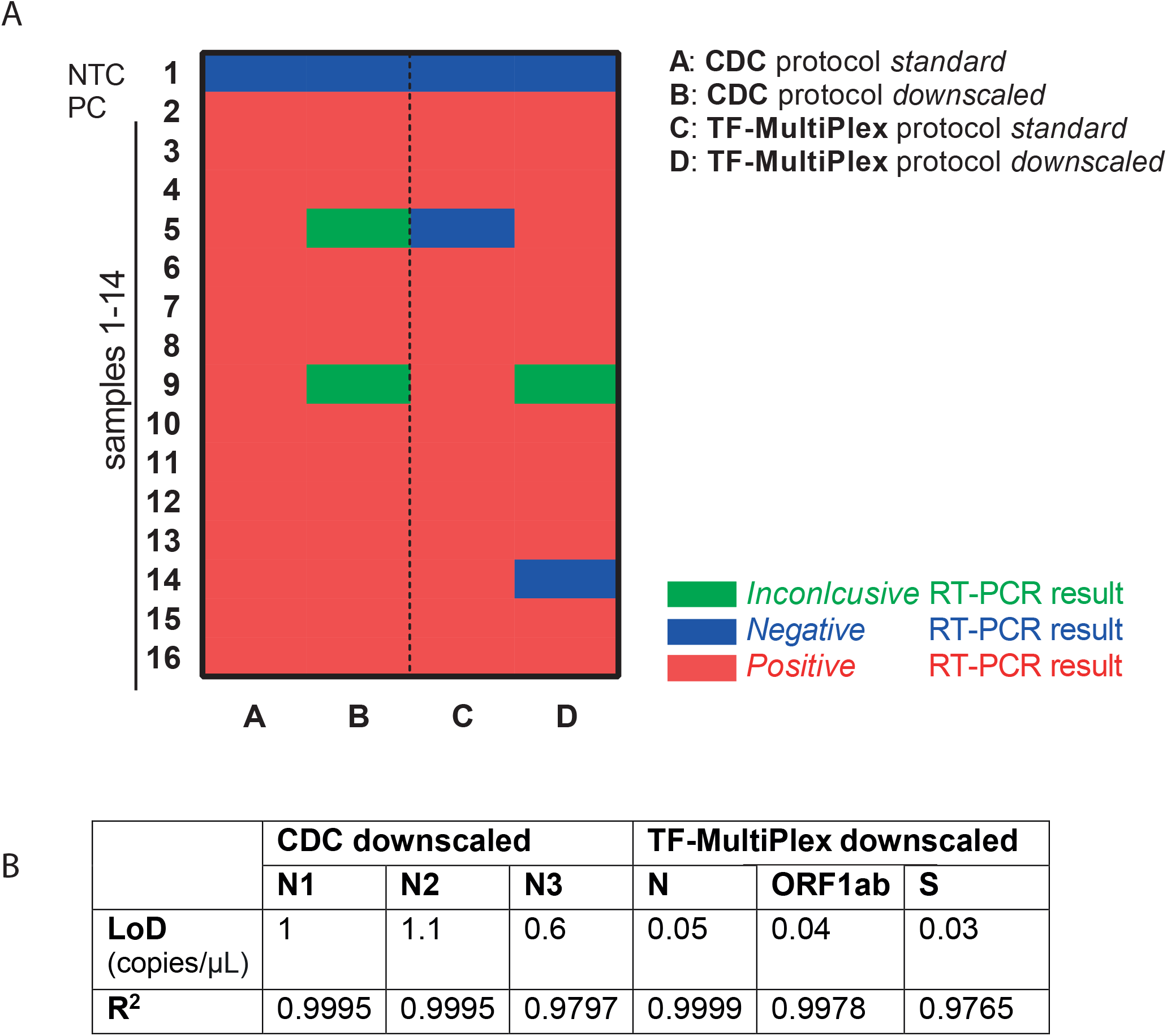
Downscaling of the CDC and TF-MultiPlex protocols. **A**. Heatmap summarizing results of standard and downscaled protocol (CDC, TF-MultiPlex); PC – positive control; NTC – Non-template control. For CDC protocol a RT-PCR result was defined as *inconclusive* if either RT-PCR was only positive for N1 (plus/minus N3) or only for N2 (plus/minus N3). For TF-MultiPlex a RT-PCR result was considered as *inconclusive* if only one of the viral genes was positive. **B**. Limit of detection (copies/µL) and R^2^ values of downscaled protocols (CDC, TF-MultiPlex).

### Environmental study of household of a SARS-CoV-2 positive patient

Having compared and established the RT-PCR protocols for SARS-CoV-2 diagnostics, we conducted a local environmental study of the presence and infectivity of SARS-CoV-2 on different surfaces in a household of a SARS-CoV-2 positive patient.

On 4^th^ of April 2020, a 42 year old female (P1, mother) patient was the first person to test positive in a family of four (Fig. 5A; Ct values at the time of diagnosis are missing, as patient was tested in a different laboratory). On 7^th^ of April 2020, the patient suffered from shortness of breath, a manifestation that lasted for three days. On the 7th of April 2020, the 42 year old male (father, P2) patient (Figure 5A) tested positive for SARS-CoV-2, symptoms resolving after 10 days. As the symptoms gradually resolved, we observed rising Ct-values in both patients (Figure 5A). On the 12^th^ of April, the two children (6 years old male (P3), 4 year old male (P4)) also tested positive for SARS-CoV-2, but remained asymptomatic at all times (Figure 5A).

**Figure 5:**
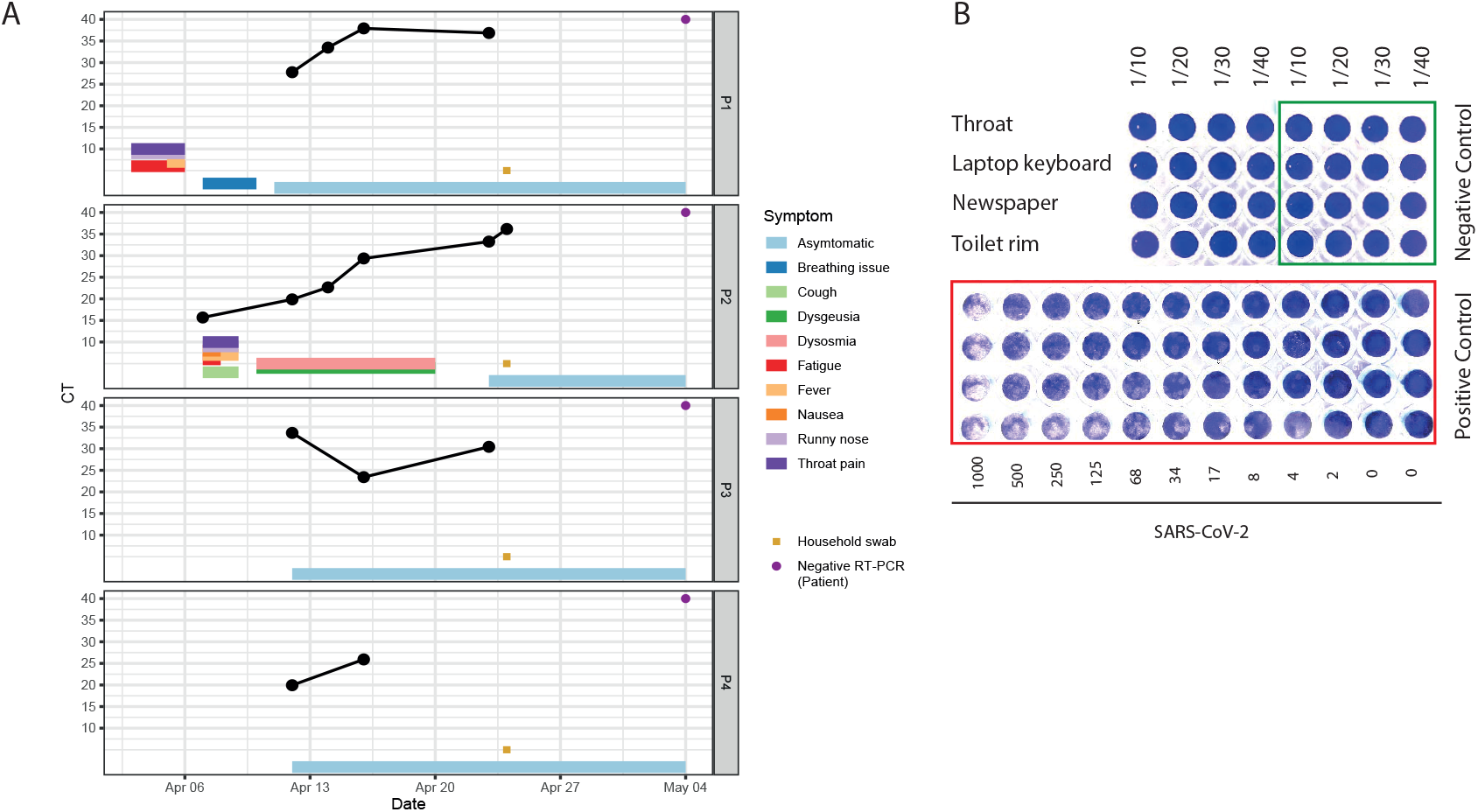
Environmental study of SARS-CoV-2 positive patient’s household. **A**. The patient’s Ct values and symptom progression. Mean of Ct values of N1, N2 and N3 viral genes are shown (CDC protocol). *P1*: 42 years old female, *P2*: 42 years old male, *P3*: 6 years old male, *P4*: 4 years old male patient. Household of the family was swabbed on the 25 of April with patients being asymptomatic for at least 2 days. **B**. An infectivity assay using Vero E6 cells showed no plaque formation for any of the samples that tested positive by real-time RT-PCR (patient’s throat, laptop keyboard, newspaper and toilet rim).

On the day of household swabbing (25^th^ of April 2020), only the P2 was swabbed again and tested positive, but reported no symptoms. The patient’s swab sample as well as surface samples were tested with three different SARS-CoV-2 detection protocols (CDC, TF-MultiPlex, and ddPCR). The laptop keyboard sample tested positive in all three SARS-CoV-2 detection protocols (S8 Table). Two more samples, showed “inconclusive” results in two independent runs with the CDC RT-PCR and in ddPCR tests, viral copies were detected from the same newspaper and a door handle, but remained below the limit of detection (S8 Table). Importantly, no infectivity was observed for the SARS-CoV-2 positive patient and the household samples (i.e., laptop keyboard, newspaper, toilet rim) (Figure 5B).

## Discussion

Real time RT-PCR remains the most sensitive method for early detection of SARS-CoV-2. Here, we report a comparison of LoD, specificity, sensitivity, economic, and practical advantages of four commercial SARS-CoV-2 detection kits as well as one optimized in house RT-PCR SARS-CoV-2 protocol.

In this study, we observed a low LoD and high sensitivity for four commercial SARS-CoV-2 RT-PCR detection protocols by using standard quantitative test samples (9) and a cohort of 92 SARS-CoV-2 positive samples, respectively. A study comparing RT-PCR to rapid fluorescence immunochromatographic (FIC) assay based SARS-CoV-2 Nucleocapsid protein (NP) antigen detection method showed that sensitivity of the rapid method was only around 75.6% (13), therefore RT-PCR remains a more sensitive detection method for SARS-CoV-2. Furthermore, we tested and confirmed specificity of those protocols with 92 samples that either had confirmed SARS-CoV-2 negative result or were collected in pre-pandemic times from patients presenting with respiratory symptoms (listed in S7 Table).

Additionally, we demonstrated that downscaling of two commercial protocols that we chose for the diagnostic routine (CDC and TF-MultiPlex), could be an option to save resources. This is especially important in times when a high demand for SARS-CoV-2 testing causes supply chain problems as occurred at the beginning of the pandemic in Europe. As an alternative strategy to optimize costs and increase testing capacity, we developed an in-house protocol in collaboration with the diagnostics company Oncobit and adapted previously published primer sequences for multiplex analysis. The customized Oncobit potocol was the least costly and fastest protocol when compared to other commercial RT-PCR protocols tested in this study.

Finally, we conducted a local environmental study by swabbing surfaces in a quarantined household of a SARS-CoV-2 positive patient. A laptop keyboard sample showed positive results in three RT-PCR detection protocols with Ct values above 30. On the day of household swabbing, the quarantined patient tested positive (cycle thresholds >30) by pharyngeal swabbing with three different protocols, while being asymptomatic for 2 days. We expected that the patient’s personal items, such as a toothbrush, should be positive for SARS-CoV-2. However, the dry condition at the time of swabbing could explain the negative result. A laptop keyboard might have a different surface and we assume that a moist environment might preserve the virus for detection, but not enough to support viral infectivity. A possible explanation for the non-infectivity of the samples could be the rather low cycle thresholds of the samples in the RT-PCR analysis, suggesting that the amount of virus might have been too low for viral isolation. Furthermore, swabbing of the households of symptomatic or pre-symptomatic SARS-CoV-2 positive individuals would be of interest to test. Further studies are needed to investigate different swabbing conditions (wet/dry) and what is the impact of freeze-thawing cycles as well as storage in viral transport medium on the detection and infectivity of a SARS-CoV-2 virus. Interestingly, the result of a quarantined household in Zurich is in line with the study of Döhla et al. in Germany where 21 households of SARS-CoV-2 positive patients were swabbed and only 3.36% (7.6% in our study) of object samples collected from household surfaces tested positive, but non-infective (14). Two other European studies (14, 15) and our study support the hypothesis that indirect environmental transmission may only play a minor role, which needs clarification in further studies.

Summarizing the comparative study, we demonstrated that most commercial and customized RT-PCR based detection protocols are highly effective at detecting viral presence in classical nasopharyngeal and/or oropharyngeal swabs and due to its high sensitivity can be applied to testing of environmental samples.

## Data Availability

Raw data (RT-PCR and ddPCR results) are available upon request.

## Acknowledgements

We thank Gaetana Restivo for help with obtaining ethical clearance for the study. We are very grateful to Jan Kaesler, Mirka Schmid, Muriel Traexler, Melanie Maudrich and the whole Dermatology biobank team of USZ for big help with running the experiments and technical help.

## Author Contribution

The idea of the study: MPL, PPB, AT, CIS, Adz, PFC.. Experimental design: MPL, PPB, AT, CIS, EB, ADz, PJ. Experiment execution: AT, CIS, EB, ADz, PJ, AD. Data analysis: CIS, PFC, AT, ADz, EB, PJ. Manuscript draft: AT, CIS. All authors edited and contributed into manuscript writing.

## Supplementary tables

**S1 Table:** Description of real-time RT-PCR assays compared in the study.

| RT-PCR protocol | Abbreviated name | RT-PCR Kit/ Primer and probes | Mastermix used in this study | Positive control |
| --- | --- | --- | --- | --- |
| CDC 2019- Novel Coronavirus Real-Time RT-PCR Diagnostic Panel (for in-vitro diagnostic | CDC | 2019-nCoV EUA-01 Diagnostic Panel Box, Cat. No. 10006606, IDT | TaMan™, Fast Virus 1-step Maste Mix, 4444436, 10mL, Applied biosystems by Thermo Fisher | 2019-nCoV_N_Positive Control, Cat. No. 10006625, IDT |
| Applied Biosystems™ TaqMan™ 2019-nCoV Assay Kit v1 | TF-SinglePlex | TaqMan™ 2019-nCoV Assay Kit v1, Cat. No. A47532, Applied biosystems by Thermo Fisher | TaMan™, Fast Virus 1-step Maste Mix, Cat. No. 4444436, 10mL, Applied biosystems by Thermo Fisher | 2019-nCoV Control v1, Cat. No. A47533, Applied biosystems by Thermo Fisher |
| Applied Biosystems™ Multiplex TaqMan 2019-nCoV Assay Kit v2 research use only (R.U.O.) kit | TF-MultiPlex | TaqPath™ COVID-19 Combo Kit, Cat. No. A47813/A47814, Applied biosystems by | TaqPath™ 1-Step Multiplex Master Mix (No ROX™) (4X) Cat. No. A28523, Applied | Positive Control (TaqPath™ COVID-19 Control Kit) Cat. No. A47816), Applied biosystems by |
|  |  | Thermo Fisher | biosystems by<br>Thermo Fisher | Thermo Fisher |
| EURORealTime<br>SARS-CoV-2<br>(For research<br>use only<br>(RUO)) | Euroimmun | Cat. No MP<br>2606-0425 | Provided with<br>the kit | Provided with the<br>kit |
| Real-time RT-PCR assays for<br>the detection of<br>SARS-CoV-2<br>Institute<br>Pasteur, Paris | Pasteur<br>Protocol<br>Pairs<br>(WHO) | (16, 17)<br>Ordered from<br>Microsynth | Invitrogen<br>Superscript™<br>III Platinum®<br>One-Step qRT-PCR system<br>Cat. No.<br>11732-088 | Available upon<br>request from<br>Pasteur Institute |
| In-house<br>customized RT-PCR protocol | Oncobit | (18, 19)<br>Ordered from<br>Microsynth | TaqPath™ 1-Step Multiplex<br>Master Mix (no<br>ROX) (A28521,<br>ThermoFisher) | SARS-CoV-2<br>Positive Run<br>Control<br>(#COV019CE) of<br>Bio-Rad |

**S2 Table:**
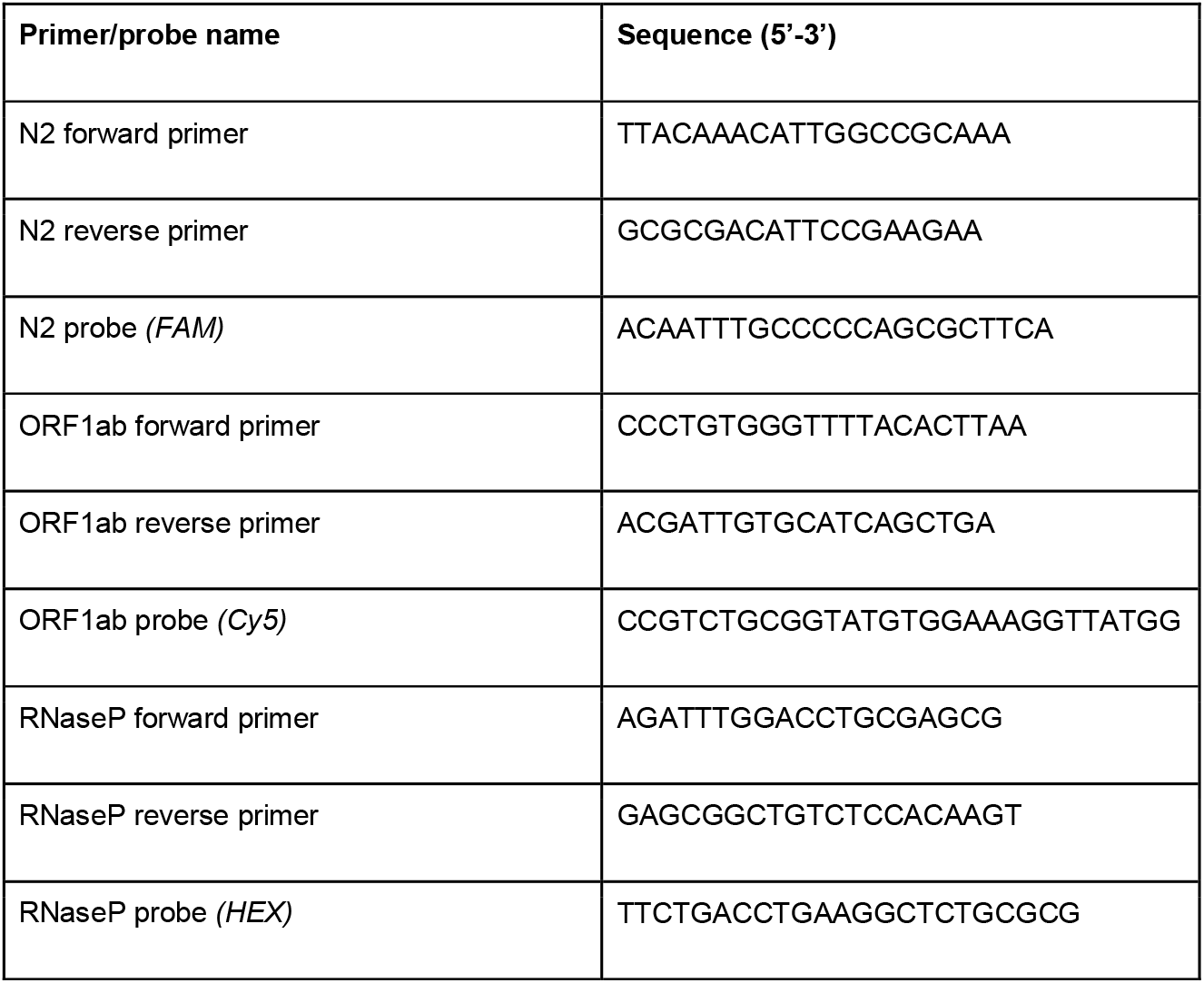
Oligonucleotide sequences of primers and probes of Oncobit real-time RT-PCR and ddPCR protocols

**S3 Table:** Reaction mix for Oncobit real-time RT-PCR protocol

| Reagents | Volume per reaction: |
| --- | --- |
| TaqPath™ 1-Step Multiplex Master Mix<br>(no ROX) (A28521, ThermoFisher) 4X | 5µL |
| N2 probe (FAM) (100µM) | 0.05µL |
| ORF1ab probe (Cy5) (100µM) | 0.05µL |
| RNaseP probe (HEX) (100µM) | 0.05µL |
| N2 forward primer (100µM) | 0.06µL |
| N2 reverse primer (100µM) | 0.06µL |
| ORF1ab forward primer (100µM) | 0.06µL |
| ORF1ab reverse primer (100µM) | 0.06µL |
| RNaseP forward primer (100µM) | 0.03µL |
| RNaseP reverse primer (100µM) | 0.03µL |
| Nuclease free H<sub>2</sub>O | 4.55µL |
| **Total** | **20.0µl** |

**S4 Table:** Thermal cycling conditions for Oncobit real-time RT-PCR protocol

| Stage | Step | Temperature | Time |
| --- | --- | --- | --- |
| Hold | UNG incubation | 25°C | 2 minutes |
| Hold | Reverse<br>Transcription | 53°C | 10 minutes |
| Hold | Activation | 95°C | 2 minutes |
| Cycling (40 cycles) | Denaturation | 95°C | 3 seconds |
|  | Anneal/Extension | 60°C | 30 seconds |

**S5 Table:** Thermal cycling conditions for Oncobit ddPCR protocol

| Stage | Temperature | Time |
| --- | --- | --- |
| Hold | 50°C | 60 minutes |
| Hold | 95°C | 10 minutes |
| Cycling (55 cycles) | 95°C | 30 seconds |
|  | 59°C | 1 minute |
| Hold | 98°C | 10 minutes |
| Hold | 4°C | 1 minute |

**S6 Table:** Results (Ct values) from two different experiments for RT-PCR protocol from Pasteur Institute, Paris

| Sample | IP2 gene |  | IP4 gene |  | E gene |  | RNase P(VIC) |  |
| --- | --- | --- | --- | --- | --- | --- | --- | --- |
| NTC | - | - | - | - | 39.2 | 35.1 | - | - |
| PC 10<sup>7</sup> copies | 20.8 | 18.2 | 19.7 | 18.9 | 18.2 | 18 | - | - |
| PC 10<sup>5</sup> copies | 30.4 | 25.5 | 28 | 27.1 | 29.2 | 25.7 | - | - |
| melanoma cell<br>line | - | - | - | - | - | 35.6 | 21.1 | 22.8 |

NTC- nuclease free water, PC – positive control

**S7 Table:**
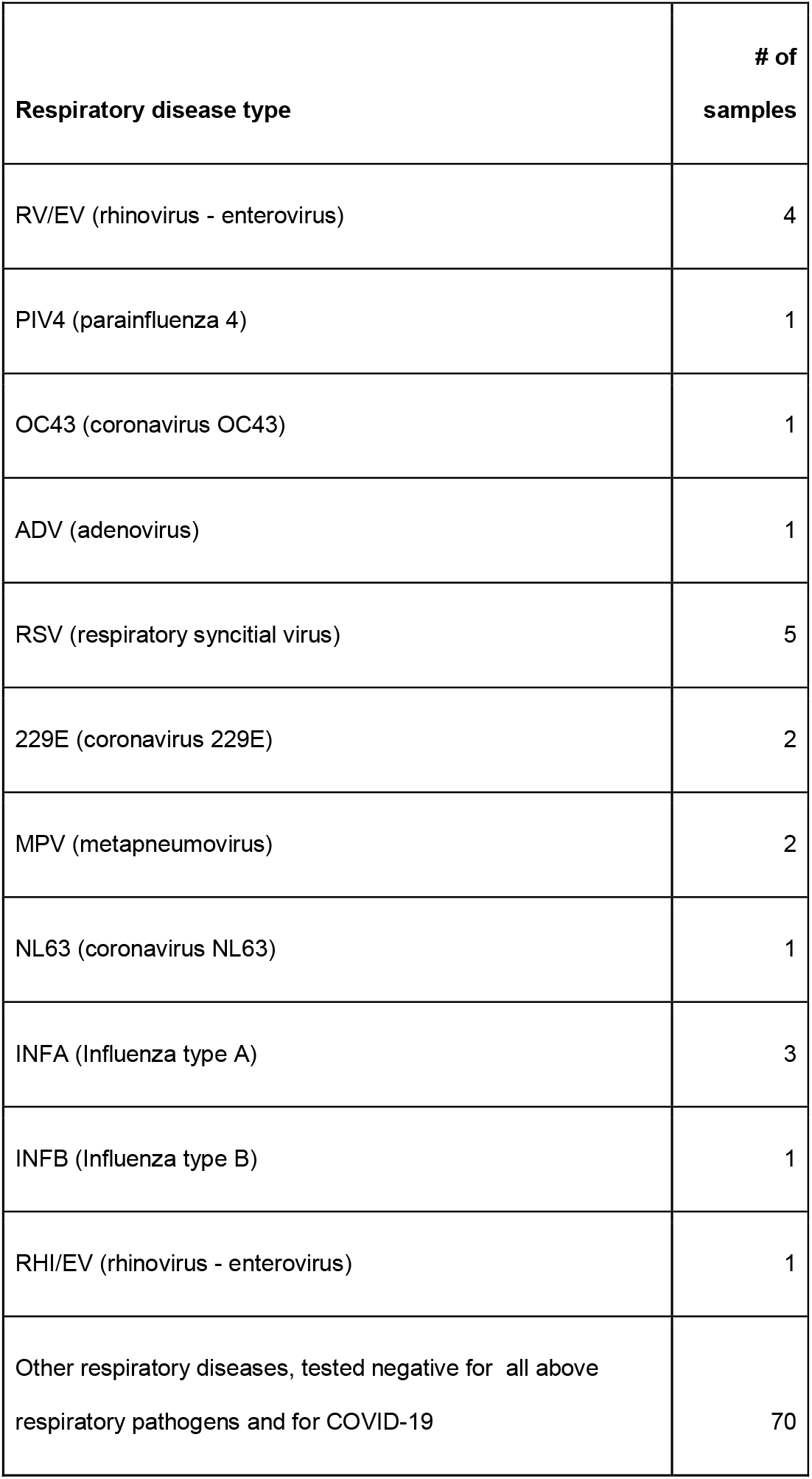
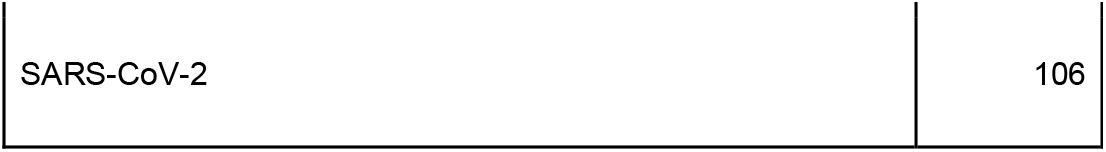
Overview of samples used in benchmarking experiments

| **Respiratory disease type** | **# of samples** |
| --- | --- |
| RV/EV (rhinovirus - enterovirus) | 4 |
| PIV4 (parainfluenza 4) | 1 |
| OC43 (coronavirus OC43) | 1 |
| ADV (adenovirus) | 1 |
| RSV (respiratory syncitial virus) | 5 |
| 229E (coronavirus 229E) | 2 |
| MPV (metapneumovirus) | 2 |
| NL63 (coronavirus NL63) | 1 |
| INFA (Influenza type A) | 3 |
| INFB (Influenza type B) | 1 |
| RHI/EV (rhinovirus - enterovirus) | 1 |
| Other respiratory diseases, tested negative for all above respiratory pathogens and for COVID-19 | 70 |

**S8 Table:**
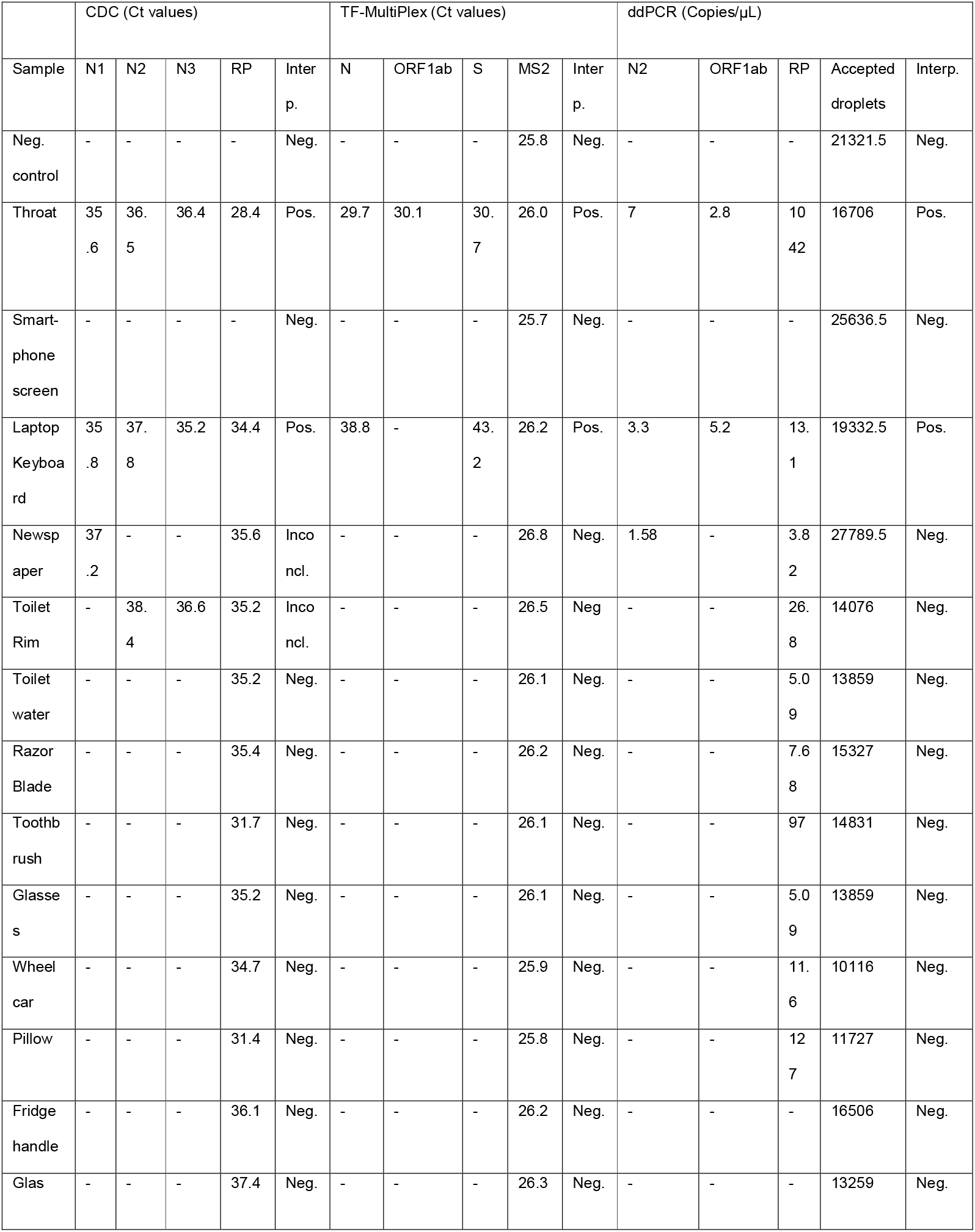
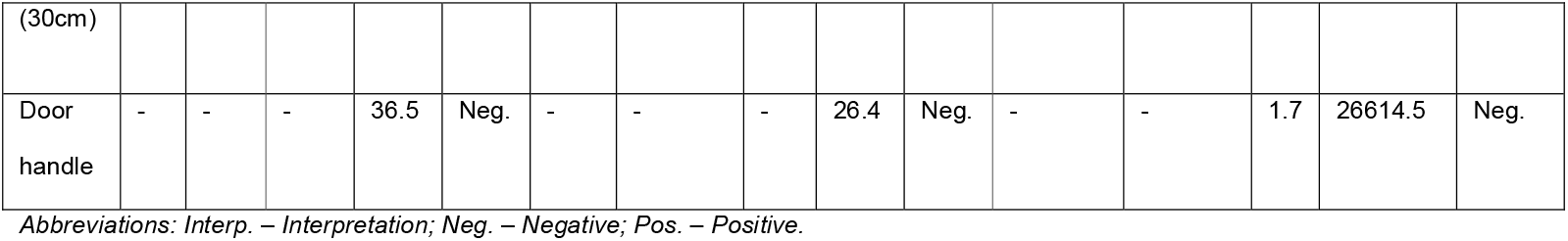
Ct values and Copies/reaction of patient’s throat and surface samples (CDC, TF-MultiPlex, ddPCR)

|  | CDC (Ct values) |  |  |  |  | TF-MultiPlex (Ct values) |  |  |  |  | ddPCR (Copies/μL) |  |  |  |  |
| --- | --- | --- | --- | --- | --- | --- | --- | --- | --- | --- | --- | --- | --- | --- | --- |
| Sample | N1 | N2 | N3 | RP | Inter<br>p. | N | ORF1ab | S | MS2 | Inter<br>p. | N2 | ORF1ab | RP | Accepted<br>droplets | Interp. |
| Neg.<br>control | - | - | - | - | Neg. | - | - | - | 25.8 | Neg. | - | - | - | 21321.5 | Neg. |
| Throat | 35<br>.6 | 36.<br>5 | 36.4 | 28.4 | Pos. | 29.7 | 30.1 | 30.<br>7 | 26.0 | Pos. | 7 | 2.8 | 10<br>42 | 16706 | Pos. |
| Smart-<br>phone<br>screen | - | - | - | - | Neg. | - | - | - | 25.7 | Neg. | - | - | - | 25636.5 | Neg. |
| Laptop<br>Keyboa<br>rd | 35<br>.8 | 37.<br>8 | 35.2 | 34.4 | Pos. | 38.8 | - | 43.<br>2 | 26.2 | Pos. | 3.3 | 5.2 | 13.<br>1 | 19332.5 | Pos. |
| Newsp<br>aper | 37<br>.2 | - | - | 35.6 | Inco<br>ncl. | - | - | - | 26.8 | Neg. | 1.58 | - | 3.8<br>2 | 27789.5 | Neg. |
| Toilet<br>Rim | - | 38.<br>4 | 36.6 | 35.2 | Inco<br>ncl. | - | - | - | 26.5 | Neg. | - | - | 26.<br>8 | 14076 | Neg. |
| Toilet<br>water | - | - | - | 35.2 | Neg. | - | - | - | 26.1 | Neg. | - | - | 5.0<br>9 | 13859 | Neg. |
| Razor<br>Blade | - | - | - | 35.4 | Neg. | - | - | - | 26.2 | Neg. | - | - | 7.6<br>8 | 15327 | Neg. |
| Toothb<br>rush | - | - | - | 31.7 | Neg. | - | - | - | 26.1 | Neg. | - | - | 97 | 14831 | Neg. |
| Glasse<br>s | - | - | - | 35.2 | Neg. | - | - | - | 26.1 | Neg. | - | - | 5.0<br>9 | 13859 | Neg. |
| Wheel<br>car | - | - | - | 34.7 | Neg. | - | - | - | 25.9 | Neg. | - | - | 11.<br>6 | 10116 | Neg. |
| Pillow | - | - | - | 31.4 | Neg. | - | - | - | 25.8 | Neg. | - | - | 12<br>7 | 11727 | Neg. |
| Fridge<br>handle | - | - | - | 36.1 | Neg. | - | - | - | 26.2 | Neg. | - | - | - | 16506 | Neg. |
| Glas | - | - | - | 37.4 | Neg. | - | - | - | 26.3 | Neg. | - | - | - | 13259 | Neg. |
| (30cm) |  |  |  |  |  |  |  |  |  |  |  |  |  |  |  |
| Door<br>handle | - | - | - | 36.5 | Neg. | - | - | - | 26.4 | Neg. | - | - | 1.7 | 26614.5 | Neg. |

Abbreviations: *Interp.* – Interpretation; *Neg.* – Negative; *Pos.* – Positive.

